# CT Attenuation Map–Derived Body Composition Is Associated with Cardiorespiratory Fitness in Multicenter External Validation

**DOI:** 10.64898/2026.05.07.26352573

**Authors:** Robert J.H. Miller, Jirong Yi, Aakash Shanbhag, Krishna Patel, Terrence D. Ruddy, Andrew J. Einstein, Attila Feher, Edward J. Miller, Joanna X. Liang, Giselle Ramirez, Leandro Slipczuk, Mark I. Travin, Erick Alexanderson, Isabel Carvajal-Juarez, René R.S. Packard, Mouaz Al-Mallah, Wanda Acampa, Stacey Knight, Viet T Le, Steve Mason, Samuel Wopperer, Panithaya Chareonthaitawee, Ronny R. Buechel, Thomas L. Rosamond, Daniel S. Berman, Damini Dey, Marcelo F. Di Carli, Piotr J. Slomka

**Affiliations:** Artificial Intelligence in Medicine Research Center, Departments of Biomedical Sciences, Medicine, and Cardiology, Cedars-Sinai Medical Center, Los Angeles, CA, United States; Department of Cardiac Sciences, University of Calgary, Calgary, AB, Canada; Signal and Image Processing Institute, Ming Hsieh Department of Electrical and Computer Engineering, University of Southern California, Los Angeles, CA, USA; Department of Medicine (Cardiology) and Population Health Science and Policy, Icahn School of Medicine at Mount Sinai, New York, NY, USA; Division of Cardiology, University of Ottawa Heart Institute, Ottawa, Ontario, Canada; Division of Cardiology, Department of Medicine, and Department of Radiology, Columbia University Irving Medical Center and New York-Presbyterian Hospital, New York, New York, United States; Section of Cardiovascular Medicine, Department of Internal Medicine, Yale University School of Medicine, New Haven, Connecticut, United States; Cardiology Division, Montefiore Health System/Albert Einstein College of Medicine, NY, USA; Department of Radiology (Nuclear Medicine), Montefiore Medical Center and Albert Einstein College of Medicine, Bronx, NY, USA; Department of Nuclear Cardiology, Ignacio Chavez National Institute of Cardiology, Mexico City, Mexico; Division of Cardiology, Department of Medicine, David Geffen School of Medicine, University of California, Los Angeles, CA, USA; Houston Methodist DeBakey Heart & Vascular Center, Houston Methodist Academic Institute, Houston, TX, USA; Department of Advanced Biomedical Sciences, University of Naples Federico II, Naples, Italy; Intermountain Medical Center Heart Institute, Intermountain Healthcare, Murray, UT, USA; Department of Cardiovascular Medicine, Mayo Clinic, Rochester, MN, USA; Department of Nuclear Medicine, Cardiac Imaging, University Hospital Zurich, Zurich, Switzerland; Department of Cardiovascular Medicine, The University of Kansas Medical Center, Kansas City, KS, USA; Cardiovascular Imaging Program, Departments of Radiology and Medicine; Division of Nuclear Medicine and Molecular Imaging, Department of Radiology; and Cardiovascular Division, Department of Medicine, Brigham and Women’s Hospital, Boston, MA, USA

**Keywords:** Deep learning, body composition, fitness, frailty

## Abstract

**Aims:** Exercise capacity is a powerful predictor of cardiovascular risk. In patients unable to exercise, body composition analysis can potentially be used to estimate cardiorespiratory fitness. We developed a body composition “fitness” score, then validated its utility in two external populations.

**Methods and Results:** We included patients from four sites undergoing single photon emission computed tomography (SPECT) and twelve sites undergoing positron emission tomography (PET). We quantified body composition using deep learning. We evaluated associations between body composition and good exercise capacity (defined as completing ≥7 minutes on a Bruce protocol) then developed a body composition “fitness” score. We then assessed the associations of “fitness” score with exercise capacity and all-cause mortality in external populations. In total, 36471 patients were included with median age 67 (interquartile range 58 – 74). Median skeletal muscle density was higher among patients with good exercise capacity. In the external SPECT population, the body composition “fitness” score had higher prediction performance for good exercise capacity (AUC 0.771, 95% CI 0.752 – 0.789) than age (AUC 0.717, p<0.01). In the external PET population, high body composition “fitness” score was associated with lower cardiovascular death (adjusted hazard ratio 0.70, 95% CI 0.62 – 0.79, p<0.001).

**Conclusions:** We demonstrated that a comprehensive body composition “fitness” score could identify patients with good cardiorespiratory fitness. This approach transforms routinely acquired CT data into a surrogate marker of fitness which can be applied in patients undergoing PET, or other CT imaging, where exercise testing is not performed.

**Graphical Abstract:** Overview of study design. The overall population (n=36471) was split as outlined above. Body composition was analyzed from available computed tomography imaging. The distribution of body composition measures were analyzed in the combined single photon emission computed tomography (SPECT) populations. A body composition “fitness” score was derived to predict good exercise capacity in the internal population, with associations assessed in the two external testing populations.

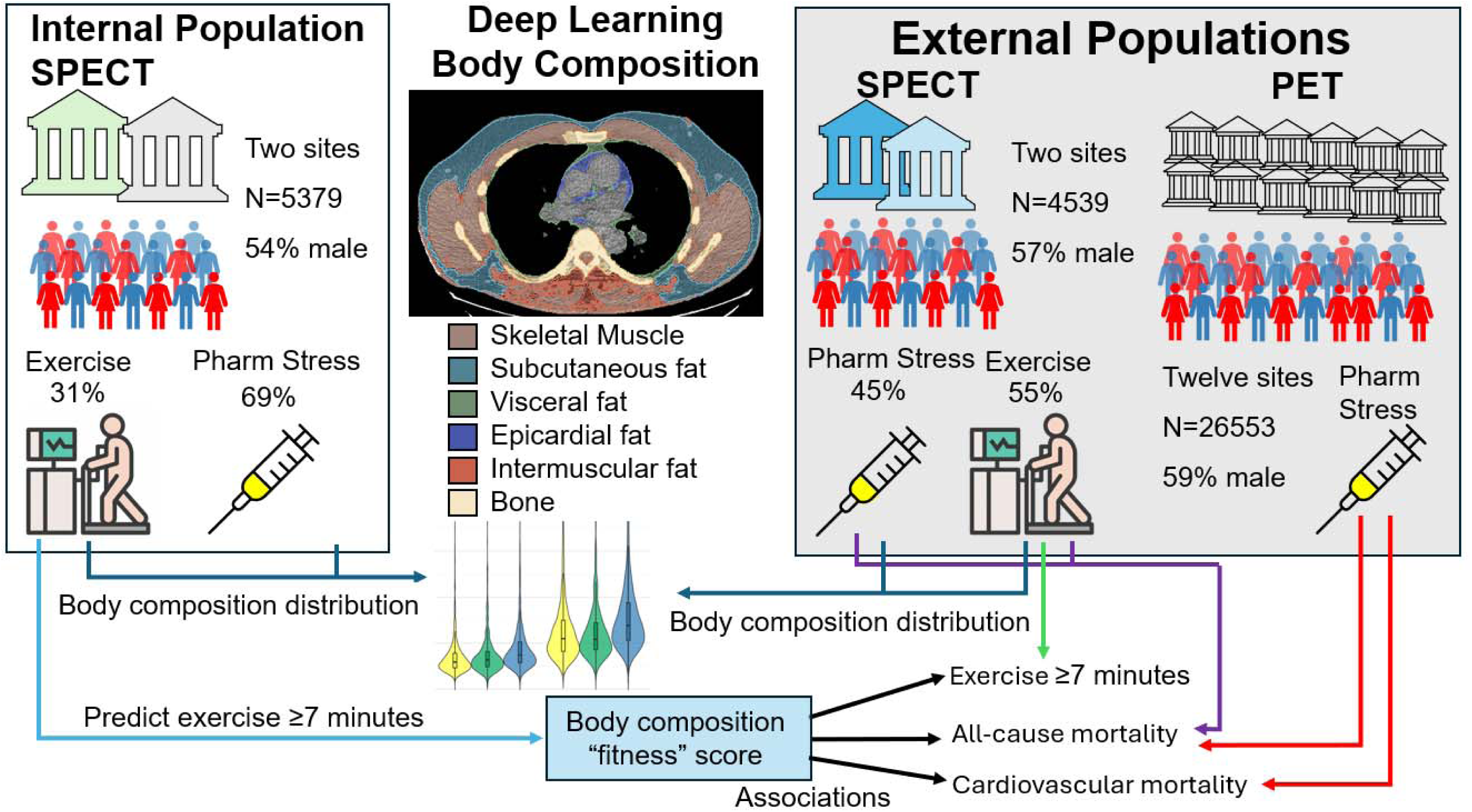

## INTRODUCTION

Frailty is a clinical syndrome characterized by decreased functional reserve and ability to tolerate stressors^1^, which is becoming increasingly common^2^. While frailty is multidimensional, many definitions focus on its physical components or “*physical frailty”*^1^. Physical frailty is inversely related to cardiorespiratory fitness^3^, which can be assessed by exercise capacity during exercise stress testing. The ability to complete exercise stress is a significant independent predictor of improved cardiovascular outcomes among patients undergoing single photon emission computed tomography (SPECT) myocardial perfusion imaging (MPI)^4^. In fact, exercise capacity is a stronger predictor of cardiovascular risk than perfusion findings in some studies^5^. However, many patients may not be able to perform exercise stress, due to medical conditions (which may be temporary), and it cannot be performed with dynamic imaging protocols typically utilized for positron emission tomography (PET).

Computed tomography (CT) scans contain a wealth of information regarding body tissue composition, which could potentially provide clinicians with critical insights into cardiorespiratory fitness. For example, skeletal muscle (SM) measurements, including volume and density, can provide quantitative measures of SM function^6, 7^. Epicardial adipose tissue (EAT), subcutaneous adipose tissue (SAT), intermuscular adipose tissue (IMAT), and visceral adipose tissue (VAT) volume and density are valuable measures of metabolic health and inflammation^8–10^. We developed a method for volumetric body composition analysis from CT attenuation correction (CTAC) images ^11, 12^. These studies highlight the potential for measuring “physical frailty” and cardiorespiratory fitness using body composition metrics.

We performed a retrospective analysis to show deep learning-based composition analysis could be used to predict cardiorespiratory fitness. We then validated the prognostic performance of a body composition “fitness” score by evaluating associations with all-cause mortality using two large external multi-center populations of patentis undergoing phramacological stress MPI.

## METHODS

### Study population

We included 9918 patients from four sites (Yale University, University of Calgary, Columbia University, University of Ottawa) who underwent SPECT/CT MPI in the REgistry of Fast Myocardial Perfusion Imaging with NExt generation SPECT (REFINE SPECT).^13, 14^ Two sites (Yale University and Ottawa Heart Institute) were used as an internal population for score derivation and two sites (University of Calgary and Columbia University) were held out for external testing. We also included an external cohort of 26553 patients undergoing PET MPI from 12 sites that were included in the REgistry of Flow and Perfusion Imaging for Artificial INtelligEnce with PET (REFINE PET)^15^. Neither Yale University nor Ottawa Heart Institute were included in the PET cohort. The study protocol complied with the Declaration of Helsinki and approval was obtained from the institutional review boards (IRB) at each participating institute, with the overall study approved at Cedars-Sinai Medical Center, Los Angeles, California.

Consent:

Patients either provided written informed consent or a waiver of consent for the use of retrospective data was granted according to site-specific protocols.

### Clinical Information

Medical history was collected at the time of clinical reporting. Prior coronary artery disease (CAD) was defined as history of myocardial infarction (MI) or previous revascularization (with percutaneous coronary intervention [PCI]) or coronary artery bypass grafting [CABG])^16^. In the SPECT MPI cohort, mode of stress was categorized as either pharmacologic stress (n=5750, 58.0%) or exercise stress (n=4168, 42.0%). The exercise stress was performed primarily using the Bruce protocol (n=3,886, 93.2%), with a smaller number of patients undergoing a modified Bruce protocol (n=282, 6.8%). Exercise stress was symptom limited, consistent with existing guideline recommendations^17^. Patients were then split based on exercise duration into patients completing at least 7 minutes and those completing less than 7 minutes. Patients who underwent a modified Bruce protocol were classified as completing less than 7 minutes. Good exercise capacity was defined as being able to complete at least 7 minutes on a Bruce protocol (which is ∼7-8 metabolic equivalents), with previous studies demonstrating that exercise capacity below this is associated with increased cardiovascular risk^18^. Patients undergoing pharmacologic stress were categorized into those undergoing pharmacologic stress with low-level exercise (n=1134, 19.7%) and those undergoing pharmacologic stress alone (n= 4616, 80.3%).

### Imaging Protocols and Quantification

SPECT MPI scanning was performed according to the American Society of Nuclear Cardiology MPI guidelines.^17^ De-identified images were transferred from each participating center to the core laboratory. Experienced core laboratory technologists conducted quality control and were blinded regarding clinical and prognostic information. Total perfusion deficit (TPD) was quantified using Quantitative Perfusion SPECT (Cedars-Sinai Medical Center, Los Angeles, CA) and used in all analyses. Left ventricular ejection fraction (LVEF) and left ventricular end-diastolic volume (LVEDV) were quantified from gated stress images using Quantitative Gated SPECT software programs (Cedars-Sinai Medical Center, Los Angeles, CA). Coronary artery calcium (CAC) was segmented and quantified automatically from CTACs with a previously validated deep learning model.^19^ PET MPI imaging was performed according to the American Society of Nuclear Cardiology MPI guidelines^20^. Quantification was performed using batch mode processing at the core laboratory as previously described^15^.

### CTAC image acquisition parameters

SPECT MPI scans were acquired with 4 different scanners as described previously ^12^. PET MPI CTAC parameters have been also described previously^15^.

### Body composition segmentation and quantification

Five body tissues (subcutaneous fat, visceral fat, bone, inter-muscular fat and skeletal muscle and bone) were quantified within T5-T11 axial coverage using our previously described hybrid approach utilizing, foundation CT models and image processing ^11, 12^. EAT was segmented separately using our previously developed deep learning model.^21, 22^ We evaluated both the volume of each tissue (indexed to height squared) and the mean attenuation (density).

### Outcomes

In the SPECT MPI population, all-cause mortality was obtained from the national death index for sites in the United States and administrative databases for sites outside of the United States. In the PET MPI population, all-cause mortality was ascertained in a similar fashion, but cause of death was also adjudicated as cardiovascular, non-cardiovascular, or unknown using standardized criteria^23^.

### Statistical analysis

Continuous variables were summarized as mean (standard deviation [SD]) if normally distributed and compared using a Student’s t-test. Continuous variables that were not normally distributed were summarized as median (interquartile range [IQR]) and compared using a Mann-Whitney U-test.

We evaluated prediction performance for good exercise capacity (as outlined above) using area under the receiver operating characteristic curve (AUC). A combined body composition “fitness” score, including volume and density of all body composition components, was created using logistic regression. Age was not included in the logistic regression model in order to avoid confounding the subsequent assessment of the association between “fitness” score and mortality, since age is a strong predictor of both fitness and mortality. The score ranges from 0 to 1 and reflects the estimated likelihood of the patient completing at least 7 minutes on a Bruce protocol. The score was developed using patients who completed exercise stress in the internal training population and then evaluated in the external testing population. The derivation population was restricted to patients undergoing exercise stress because exercise capacity was known. Optimal thresholds were derived using the Youden index in the internal population.

In order to validate the utility of the “fitness” score, we evaluated associations with all-cause mortality using univariable and multivariable Cox proportional hazards analyses. Multivariable models in the SPECT MPI population included age, sex, body mass index (BMI), medical history (hypertension, diabetes, dyslipidemia, prior CAD), smoking history, family history of CAD, stress TPD, LVEF, and CAC. Multivariable models in the PET MPI population also included LVEF reserve and myocardial flow reserve (MFR). All statistical tests were two-sided, and a p-value <0.05 was considered statistically significant. All analyses were performed using Stata/IC version 13.1 (StataCorp, College Station, Texas, USA) and R (version 4.1.2).

## RESULTS

### Study Population

In total, 36471 patients were included with median age 67 (interquartile range 58 – 74) and 21162 (58.0%) male patients. A comparison of the internal and external testing populations is shown in **Table 1**. Patients undergoing PET MPI were older (median age 67 vs 64 and 66, p<0.001) and had a higher prevalence of all comorbidities.

**Table 1:** Population characteristics of the internal and external single photon emission populations. BMI - body mass index, CAD - coronary artery disease, IQR - interquartile range

|  | Overall<br>(N=36471) | Internal SPECT<br>n=5379 | External SPECT<br>n=4539 | External PET<br>N=26553 | p-value |
| --- | --- | --- | --- | --- | --- |
| Age, median (IQR) | 67 (58, 74) | 64 (56, 73) | 66 (58, 74) | 67 (58, 75) | <0.001 |
| Male, n (%) | 21162 (58.0%) | 2885 (53.6%) | 2566 (56.5%) | 15711 (59.2%) | 0.004 |
| BMI, median (IQR) | 29.7 (25.7, 34.9) | 30.7 (26.6, 34.9) | 29.6 (25.7, 34.4) | 29.5 (25.6, 34.9) | <0.001 |
| Past medical history, n (%) |  |  |  |  |  |
| Hypertension | 27003 (74.3%) | 3508 (65.2%) | 2659 (58.6%) | 20836 (78.9%) | <0.001 |
| Diabetes mellitus | 12601 (34.6%) | 1450 (27.0%) | 1372 (30.2%) | 9779 (36.9%) | <0.001 |
| Dyslipidemia | 24341 (67.0%) | 2912 (54.1%) | 2146 (47.3%) | 19283 (73.0%) | <0.001 |
| Family history | 9269 (25.7%) | 1031 (19.2%) | 457 (10.1%) | 6807 (26.0%) | <0.001 |
| Smoking | 6328 (17.4%) | 1173 (21.8%) | 1138 (25.1%) | 4840 (18.3%) | <0.001 |
| Prior CAD | 10552 (28.9%) | 1173 (21.8%) | 1138 (25.1%) | 8241 (31.0%) | <0.001 |
| Stress Testing |  |  |  |  | <0.001 |
| Pharmacologic | 32303 (88.6%) | 3723 (69.2%) | 2027 (44.7%) | 26553 (100%) |  |
| Exercise < 7 minutes | 1858 (5.1%) | 498 (9.3%) | 1360 (30.0%) | 0 (0%) |  |
| Exercise ≥ 7 minutes | 2310 (6.3%) | 1158 (21.5%) | 1152 (25.4%) | 0 (0%) |  |

Characteristics of the SPECT MPI population stratified by mode of stress are shown in **Table 2**. Patients undergoing pharmacologic stress were older (median age 67 vs 62, p<0.0001) and had a higher BMI (30.9 vs 29.2, p<0.001). Patients undergoing pharmacologic stress also had higher rates of cardiovascular risk factors (hypertension 67.7% vs 54.6%, p<0.001; diabetes 32.8% vs 22.4%, p<0.001) and history of prior CAD (25.7% vs 20.1%, p<0.001).

**Table 2:** Population characteristics for the single photon emission computed tomography patients split by stress testing modality and exercise duration. BMI - body mass index, CAD - coronary artery disease, IQR - interquartile range

|  | Overall<br>SPECT<br>(N=9918) | Pharmacologic Stress<br>SPECT<br>n=5750 | Exercise Stress<br><7 minutes<br>n=1858 | Exercise Stress<br>≥7 minutes<br>n=2310 | p-value |
| --- | --- | --- | --- | --- | --- |
| Age, median (IQR) | 65 (57, 73) | 67 (59, 75) | 67 (59, 75) | 59 (51, 65) | <0.001 |
| Male, n (%) | 5451 (55.0%) | 2948 (51.3%) | 896 (48.2%) | 1607 (69.6%) | <0.001 |
| BMI, median (IQR) | 30.1 (26.1, 34.7) | 30.9 (26.6, 35.5) | 30.5 (26.3, 35.2) | 28.5 (25.4, 32.3) | <0.001 |
| Past medical history, n (%) |  |  |  |  |  |
| Hypertension | 6167 (62.2%) | 3891 (67.7%) | 1159 (62.4%) | 1117 (48.4%) | <0.001 |
| Diabetes mellitus | 2822 (28.5%) | 1887 (32.8%) | 521 (28.0%) | 414 (17.9%) | <0.001 |
| Dyslipidemia | 5058 (51.0%) | 3079 (53.5%) | 908 (48.9%) | 1071 (46.4%) | <0.001 |
| Family history | 2462 (24.8%) | 984 (17.1%) | 210 (11.3%) | 294 (12.7%) | <0.001 |
| Smoking | 1488 (15.0%) | 1475 (25.7%) | 370 (19.9%) | 466 (20.2%) | <0.001 |
| Prior CAD | 2311 (23.3%) | 3891 (67.7%) | 1159 (62.4%) | 1117 (48.4%) | <0.001 |

### Body Composition by Mode of Stress and Exercise Capacity

Skeletal muscle volume and attenuation stratified by sex and stress are shown in **Figure 1**. Male patients who were able to exercise for at least 7 minutes had significantly higher skeletal muscle volume and attenuation compared to male patients who underwent pharmacologic stress (volume median 968 vs 878, p<0.001; attenuation median 73 vs 31, p<0.001). Similar results were seen for female patients (median volume 690 vs 663, p<0.001; median attenuation 31 vs 25, p<0.001). Total muscle to adipose tissue ratio stratified by sex and stress is shown in **Figure 2**. Among female patients, muscle to adipose tissue was highest in patients able to complete at least 7 minutes (median 0.39), followed by patients exercise less than 7 minutes (median 0.32) and patients requiring pharmacologic stress (median 0.29, p<0.001 for all comparisons). Among male patients, median muscle to adipose ratio was significantly higher in patients undergoing exercise for at least 7 minutes (median 0.72) compared to patients exercising less than 7 minutes (median 0.56, p<0.001) or requiring pharmacologic stress (median 0.55, p<0.001). A summary of all body composition metrics stratified by sex and stress is shown in **Supplemental Table 1.**

**Figure 1:**
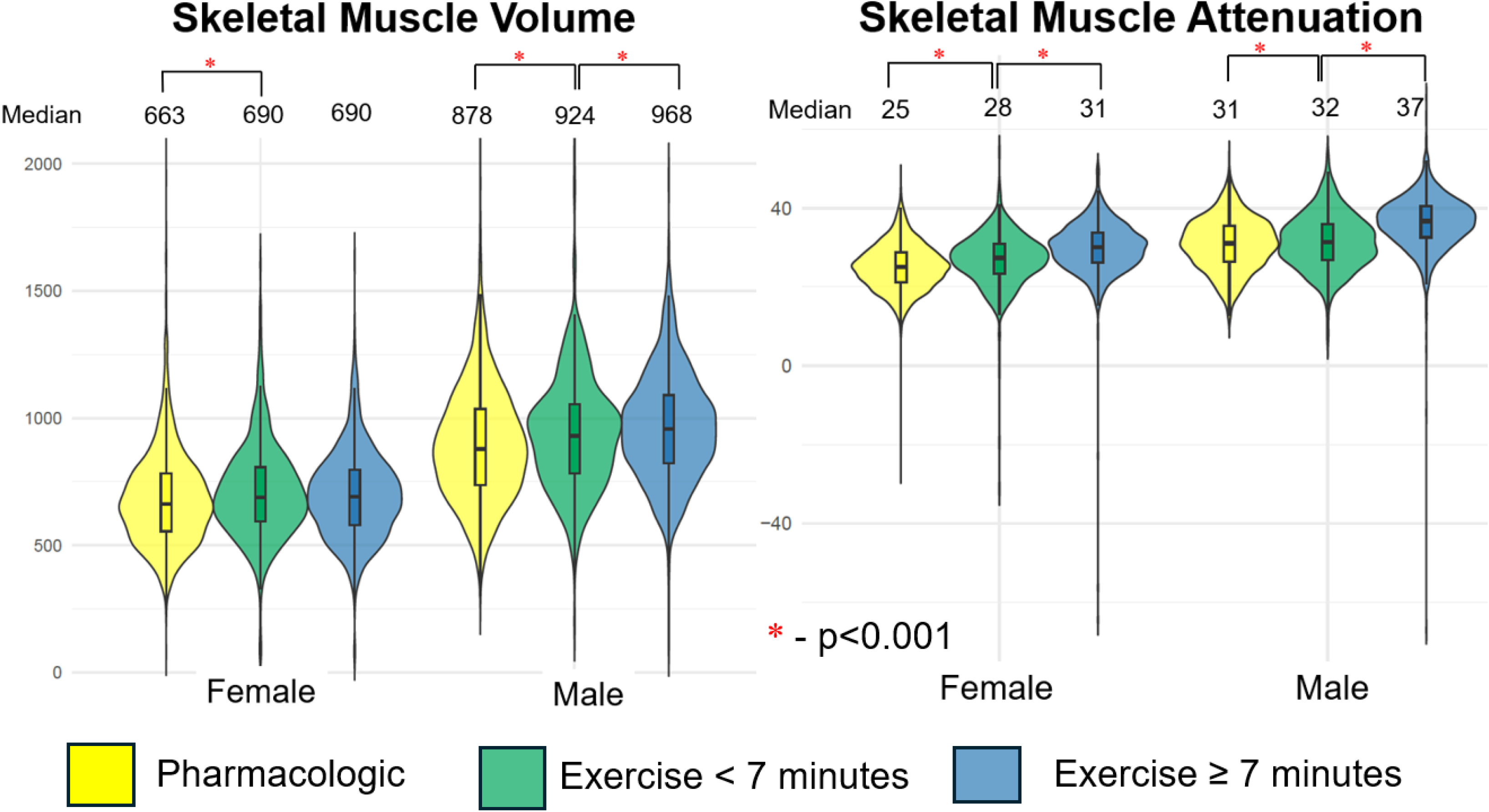
Skeletal muscle volume and attenuation stratified by mode of stress. Box plots demonstrate median as well as 25^th^ and 75^th^ percentile with superimposed violin plots outlining distribution of all values.

**Figure 2:**
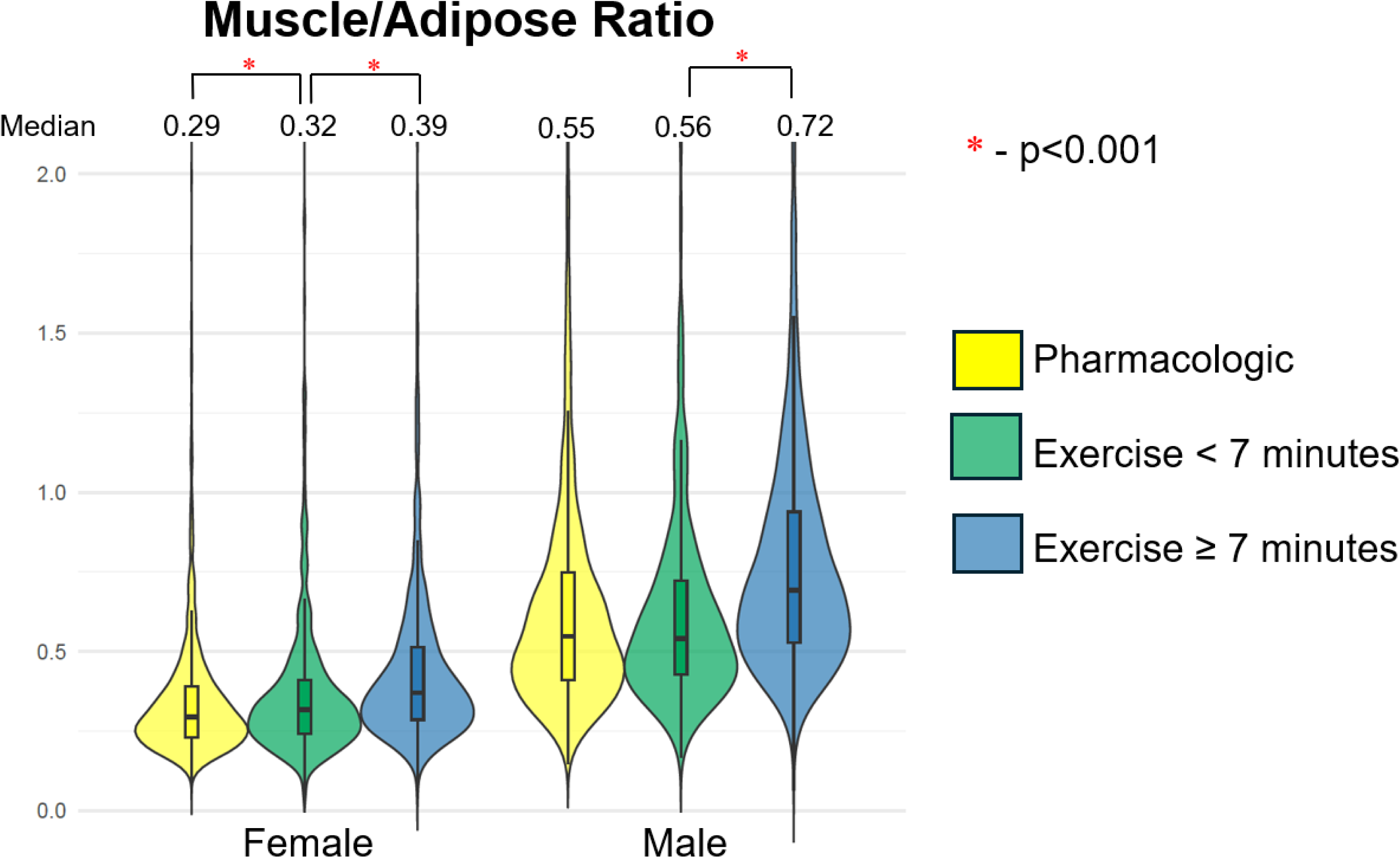
Muscle to adipose tissue ratios stratified by sex and mode of stress. Box plots demonstrate median as well as 25^th^ and 75^th^ percentile values with superimposed violin plots outlining distribution of all values.

### Predicting Cardio-Respiratory Fitness

Body composition metrics were combined using logistic regression, in the internal SPECT population, separately by sex (models shown in **Table 3**). In female patients, SM attenuation (OR 1.105, p<0.001), VAT volume index (OR 0.997, 95% CI 0.021), IMAT volume index (OR 1.006, p=0.002) and bone attenuation (OR 1.006, p=0.003) were associated with good exercise capacity. In male patients, SM volume index (OR 1.002, p=0.024), SM attenuation (OR 1.073, p=0.005), SAT volume index (OR 0.999, p=0.042), VAT attenuation (OR 0.926, p=0.002), and bone volume index (OR 0.995, p=0.043) were associated with good exercise capacity.

**Table 3:** Logistic regression models for predicting exercise time of at least 7 minutes among patients undergoing exercise stress. Total adipose tissue excluded due to collinearity. Model developed in the internal population. Significant associations (p<0.05) in bold. CI – confidence interval, OR – odds ratio

|  | Female |  | Male |  |
| --- | --- | --- | --- | --- |
|  | OR (95% CI) | p-value | OR (95% CI) | p-value |
| SM volume index | 0.999 (0.997 - 1.001) | 0.197 | **1.002 (1.000 - 1.003)** | **0.024** |
| SM attenuation | **1.105 (1.056 - 1.157)** | **<0.001** | **1.073 (1.021 - 1.127)** | **0.005** |
| SAT volume index | 1.000 (0.999 – 1.000) | 0.232 | **0.999 (0.999 - 1.000)** | **0.043** |
| SAT attenuation | 0.984 (0.930 - 1.040) | 0.557 | 0.972 (0.931 - 1.015) | 0.198 |
| VAT volume index | **0.997 (0.994 - 0.999)** | **0.021** | 0.999 (0.997 - 1.000) | 0.114 |
| VAT attenuation | 0.986 (0.936 - 1.040) | 0.615 | **0.926 (0.883 - 0.972)** | **0.002** |
| EAT volume index | 1.005 (0.995 - 1.014) | 0.361 | 0.991 (0.981 - 1.001) | 0.08 |
| EAT attenuation | 1.001 (0.996 - 1.005) | 0.813 | 1.002 (0.998 - 1.007) | 0.314 |
| IMAT volume index | **1.006 (1.002 - 1.010)** | **0.002** | 1.000 (0.998 - 1.003) | 0.745 |
| IMAT attenuation | 1.043 (0.999 - 1.090) | 0.058 | 1.033 (0.990 - 1.078) | 0.139 |
| Muscle to adipose ratio | 1.814 (0.602 - 5.465) | 0.290 | 1.122 (0.707 - 1.780) | 0.625 |
| Bone volume index | 1.001 (0.994 - 1.007) | 0.796 | **0.995 (0.991 – 1.000)** | **0.043** |
| Bone attenuation | **1.006 (1.002 - 1.011)** | **0.003** | 1.000 (0.99 – 1.000) | 0.468 |

The combined body composition “fitness” score ranges from 0 to 1 and reflects the predicted probability for the patient to be able to complete at least 7 minutes of exercise stress. The optimal thresholds for identifying “fit” patients were 0.769 in male patients and 0.508 in female patients, with scores above these thresholds associated with a higher likelihood of good exercise capacity. The optimal thresholds for SM attenuation were 27.4 HU and 35.3 HU for female and male patients respectively.

Prediction performance for good exercise capacity, defined as completing at least 7 minutes on a Bruce protocol, among patients completing exercise stress is shown in **Supplemental Table 2.** In the external SPECT population, the body composition “fitness” score had higher prediction performance for good exercise capacity (AUC 0.771, 95% CI 0.752 – 0.789) compared to age (AUC 0.717, 95% CI 0.698 – 0.737, p<0.01), and BMI (AUC 0.583, 95% CI 0.561 – 0.605, p<0.01). AUC could be further increased by incorporating age with the body composition score (AUC 0.804, 95% CI 0.787 – 0.821, p<0.01).

### Associations with Outcomes – External PET population

During median follow-up of 4.3 years (IQR 2.1 – 5.4), 5157 patients died (19.4%), with 2386 of those deaths classified as cardiovascular (46.3%). Details of the PET MPI population, stratified by body composition analysis, are shown in **Supplemental Table 3.** Patients categorized as “fit” by body composition analysis were at lower risk of cardiovascular death (unadjusted HR 0.50, 95% CI 0.45 – 0.56), as shown in **Figure 3**. The association with reduced cardiovascular mortality persisted after adjustment for age, sex, BMI, medical history, stress and rest TPD, stress LVEF, LVEF reserve, CAC, and MFR (adjusted HR 0.70, 95% CI 0.62 – 0.79, p<0.001). Similarly, Patients categorized as “fit” by body composition analysis were at lower risk of all-cause death in both unadjusted (HR 0.56, 95% CI 0.51 – 0.60) and adjusted analyses (HR 0.75, 95% CI 0.69 – 0.82), as shown in **Figure 4**.

**Figure 3:**
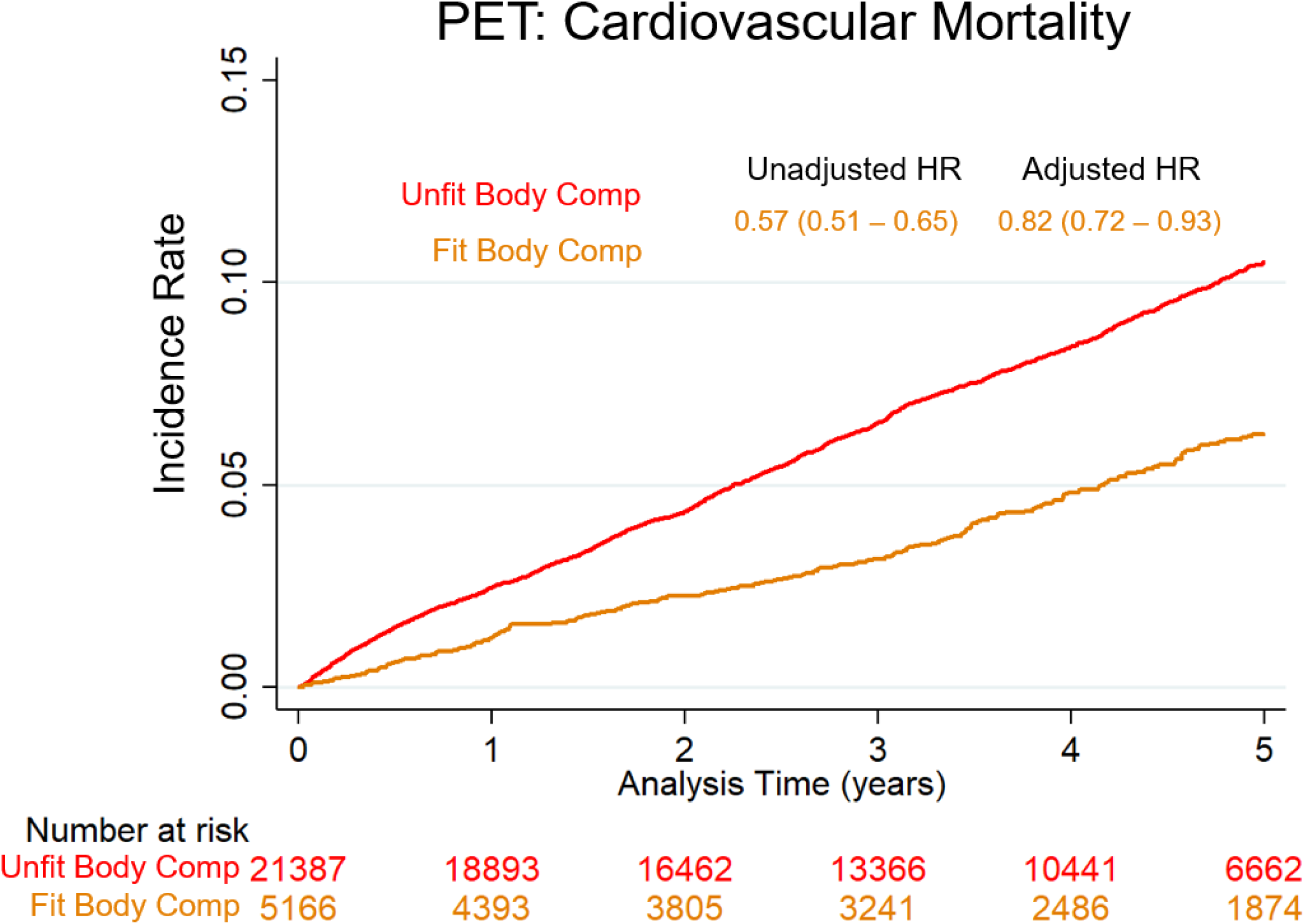
Kaplan-Meier incidence curves for cardiovascular mortality in the positron emission tomography cohort. Hazard ratios (HR) reflect the risk associated with high body composition “fitness” score. Multivariable models include age, sex, medical history, stress total perfusion deficit, stress left ventricular ejection fraction, ejection fraction reserve, coronary artery calcium, and myocardial flow reserve.

**Figure 4:**
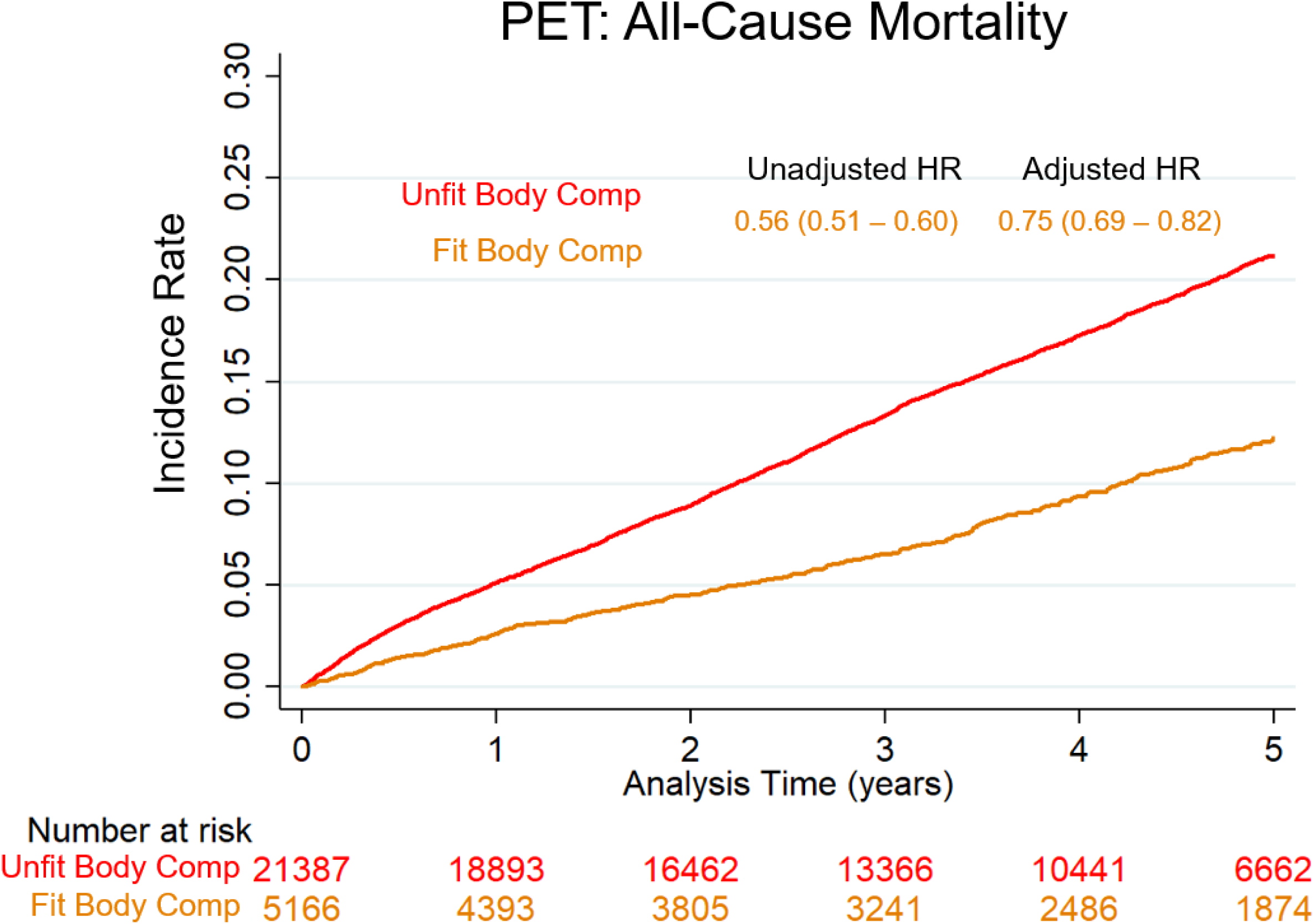
Kaplan-Meier incidence curves for all-cause mortality in the positron emission tomography cohort. Hazard ratios (HR) reflect the risk associated with high body composition “fitness” score. Multivariable models include age, sex, medical history, stress total perfusion deficit, stress left ventricular ejection fraction, ejection fraction reserve, coronary artery calcium, and myocardial flow reserve.

### Associations with All-Cause Mortality – External SPECT population

During median follow-up of 2.5 years (IQR 1.5 – 3.7), 610 (6.2%) patients died. Patients requiring pharmacologic stress were more likely to die during follow-up (8.1% vs 3.5%, p<0.001). Kaplan-Meier survival curves stratified by the need for pharmacologic stress and body composition “fitness” score in the external SPECT population are shown in **Supplemental Figure 1.** Patients who were classified as “fit” by body composition analysis were at lower risk among patients who underwent exercise stress (unadjusted HR 0.49, 95% CI 0.34 – 0.73, p<0.001; adjusted HR 0.48, 95% CI 0.31 – 0.75, p<0.001). Furthermore, “fit” body composition continued to be associated with reduced all-cause mortality after also adjusting for exercise duration (adjusted HR 0.55, 95% CI 0.33 – 0.92, p=0.024).

### Case Examples

Two cases demonstrating combined body composition “fitness” analysis are shown in **Figure 5**. In panel A, CT images and body composition segmentations from a male patient who underwent exercise stress for unstable angina and completed 3 minutes on a Bruce protocol. He had low SM volume index (458 ml/m^2^) and SM density (mean 16.5 HU). The patient died ∼3 years later. In panel B, CT images and body composition segmentation from a male patient who underwent pharmacologic stress testing for suspected acute coronary syndrome. SM volume index (1349 ml/m^2^) and SM density (mean 42.7 HU) were high. The patient has not had any cardiovascular events during > 7 years follow-up.

**Figure 5:**
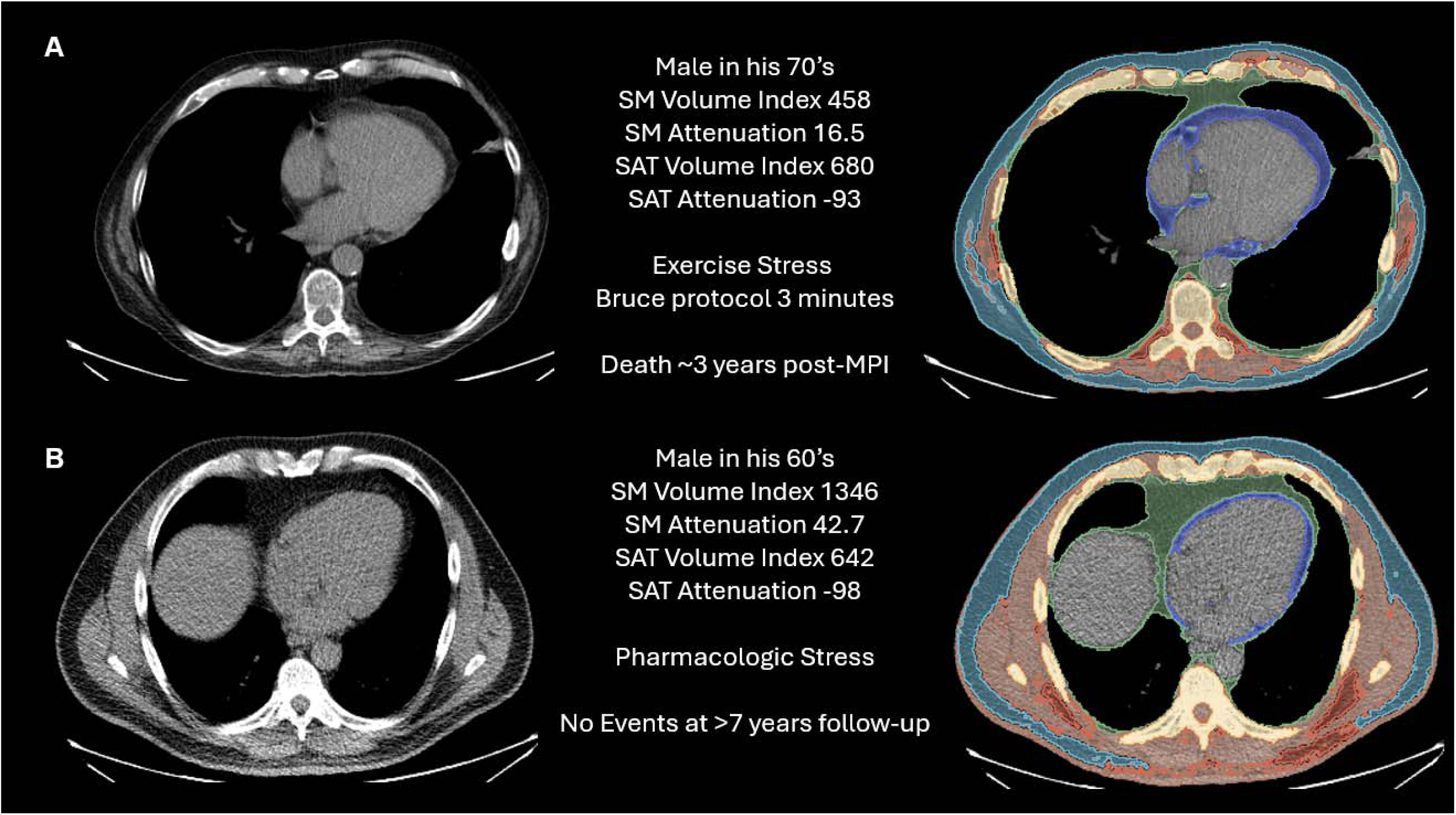
Case examples. In panel A, CT images and body composition segmentations from a male patient who underwent exercise stress for unstable angina and completed 3 minutes on a Bruce protocol. He had low SM volume index (458 ml/m2) and SM density (mean 16.5 HU). The patient died ∼ 3 years later. In panel B, CT images and body composition segmentation from a male patient who underwent pharmacologic stress testing for suspected acute coronary syndrome. SM volume index (1349 ml/m2) and SM density (mean 42.7 HU) were high. The patient has not had any cardiovascular events during > 7 years follow-up

## DISCUSSION

We utilized data from two large, multicenter registries to evaluate a method for predicting cardiorespiratory fitness from chest CT attenuation scan for patients who undergo pharmacological stress MPI. Body composition measures differed significantly between patients with good exercise capacity compared to those with lower exercise capacity or requiring pharmacologic stress. We integrated SM, adipose tissue, and bone measures to create a comprehensive “fitness” score which was better able to predict exercise capacity compared to variables such as age and BMI. We went on to validate the utility of these measures by demonstrating associations with both all-cause and cardiovascular mortality in a large, multicenter cohort of patients undergoing PET MPI. Our results suggest that a deep learning-based fitness score could be used to identify patients with good exercise capacity, providing valuable clinical insights.

One strength of SPECT MPI is the ability to provide a comprehensive clinical evaluation including exercise capacity. While exercise capacity is a valuable marker of cardiovascular risk^18^, not all patients are able to complete treadmill stress. For example, patients with orthopedic injuries or awaiting orthopedic surgeries may not be able to complete treadmill stress, but this does not directly reflect poor cardiorespiratory fitness. We recently demonstrated that requirement for pharmacologic stress testing was not associated with increased risk in patients undergoing SPECT MPI for pre-operative indications^24^, presumably due to transient conditions precluding exercise stress. Additionally, exercise stress cannot be combined with standard myocardial blood flow studies due to the need for early dynamic imaging^25^. Furthermore, exercise capacity is not measured with coronary CT angiography or coronary artery calcium scoring but our approach could potentially be applied in these populations. Lastly, even among patients who could exercise, the body composition “fitness” score provided independent risk stratification after adjusting for relevant confounders including exercise duration. Our proposed “fitness” score, based on comprehensive body composition analysis, could provide new insights into the cardiorespiratory health for a broad range of patients undergoing cardiovascular testing.

Among patients undergoing exercise stress, we identified significant differences in body composition between patients who completed at least 7 minutes on a Bruce protocol compared to those who could not. In particular, we identified significant differences in SM attenuation in both male and female patients as well as significant difference in SM volume in male patients. Both SM attenuation and volume are associated with muscle strength^6, 7^. As such, both measures can be considered markers of sarcopenia - a progressive and generalized disorder characterized by loss of skeletal muscle function^26–28^. Conservative estimates suggest that 5 to 10% of the general population could be considered sarcopenic^28^. However, patients with cardiovascular disease have a higher prevalence of sarcopenia compared to the general population, with rates ranging from 15% among patients with congenital heart disease to 32% among patients with heart failure^29^. Sarcopenia is associated with physical disability^26, 27^ and mortality in patients with heart failure and other cardiovascular disorders^27, 30^. Importantly, supervised exercise programs for patients with sarcopenia can improve muscle mass and reverse the associated physical frailty^31^. Therefore, the proposed body composition analysis could not only be used to identify patients with poor cardiorespiratory fitness, but it could also potentially help target interventions to improve patient outcomes.

Our results are consistent with other studies in the literature evaluating the utility of body composition analysis to predict cardiorespiratory fitness. Heileson et al. demonstrated that percent body fat and bone mineral content, measured with dual-energy x-ray absorptiometry, was associated with maximal oxygen consumption in a cohort of 60 patients undergoing cardiopulmonary exercise testing^32^. Similarly, Marks et al. found that percent body fat was associated with fitness measured using a shuttle run test among 375 adolescents^33^. In our analysis, muscle to adipose tissue ratio (analogous to body fat percentage) and bone metrics were associated with good exercise capacity. Furthermore, SM attenuation (which cannot be measured with dual-energy x-ray absorptiometry) was a stronger predictor suggesting benefits from utilizing existing CT information. Importantly, we demonstrated these findings in a substantially larger population from an international, multicenter registry. Additionally, we provided further validation by demonstrating that the body composition fitness score predicted risk of all-cause or cardiovascular mortality in patients undergoing PET MPI. This is a population where this approach has greatest potential, since the dynamic acquisitions require for myocardial blood flow measurements cannot be combined with exercise stress. However, it could also potentially be applied to other chest CT imaging, such as coronary CT angiography or CAC scans.

The present study has a few important limitations. We relied on fully automated body composition segmentations, which may not completely agree with expert manual segmentations. However, given the excessive time required for manual segmentations they are not likely to be feasible clinically. We had a limited definition of cardiorespiratory fitness, based only on exercise duration on treadmill stress. Duration of treadmill exercise stress is a strong predictor of all-cause mortality risk^34^, but does not capture all aspects of cardiorespiratory fitness or physical frailty.

## CONCLUSIONS

We demonstrated that a comprehensive body composition CT-based “fitness” score could identify patients with good exercise capacity, and higher scores were associated with a reduced risk of both all-cause and cardiovascular mortality in a large external population. This approach transforms routinely acquired CT data into a surrogate marker of fitness which can be applied in patients undergoing PET MPI or other CT imaging where exercise testing is not routinely performed.

## DATA AVAILABILITY STATEMENT

The data underlying this manuscript can be made available following written request to the corresponding author and after appropriate data sharing agreements are in place.

## ACKNOWLEDGEMENTS

None

