## Supplemental Material for "CT Attenuation Map–Derived Body Composition Is Associated with Cardiorespiratory Fitness in Multicenter External Validation"

| Female Patients (n=4467) | Pharmacologic Stress | Exercise < 7 minutes | Exercise ≥ 7 minutes | p-values |
| --- | --- | --- | --- | --- |
|  | n=2802 | n=962 | n=703 |  |
| SM volume index | 663 (555, 782) | 690 (588, 808) | 690 (578, 793) | <0.001 |
| SM attenuation | 25 (21, 29) | 28 (24, 31) | 31 (27, 35) | <0.001 |
| SAT volume index | 1792 (1249, 2372) | 1759 (1194, 2334) | 1321 (883, 1857) | <0.001 |
| SAT attenuation | -103 (-106, -99) | -103 (-105, -99) | -103 (-105, -99) | 0.004 |
| VAT volume index | 324 (224, 447) | 325 (222, 443) | 233 (157, 329) | <0.001 |
| VAT attenuation | -82 (-86, -78) | -83 (-87, -79) | -81 (-85, -77) | <0.001 |
| EAT volume index | 49 (35, 68) | 48 (34, 64) | 42 (31, 57) | <0.001 |
| EAT attenuation | -63 (-70, -54) | -59 (-66, -54) | -55 (-62, -50) | <0.001 |
| IMAT volume index | 79 (54, 115) | 72 (49, 111) | 58 (38, 89) | <0.001 |
| IMAT attenuation | -69 (-73, -66) | -70 (-73, -66) | -69 (-72, -66) | 0.21 |
| Total adipose tissue | 2287 (1643, 2978) | 2270 (1580, 2901) | 1689 (1172, 2332) | <0.001 |
| Muscle to adipose ratio | 0.29 (0.23, 0.39) | 0.32 (0.25, 0.42) | 0.39 (0.31, 0.55) | <0.001 |
| Bone volume index | 278 (250, 308) | 283 (254, 311) | 268 (240, 290) | <0.001 |
| Bone attenuation | 249 (221, 283) | 250 (219, 284) | 274 (243, 309) | <0.001 |
| Male Patients (n=5451) | Pharmacologic Stress | Exercise < 7 minutes | Exercise ≥ 7 minutes | p-values |
|  | n=2948 | n=896 | n=1607 |  |
| SM volume index | 878 (736, 1036) | 924 (793, 1057) | 968 (827, 1103) | <0.001 |
| SM attenuation | 31 (26, 35) | 32 (28, 37) | 37 (34, 41) | <0.001 |
| SAT volume index | 926 (609, 1350) | 947 (631, 1335) | 738 (521, 1022) | <0.001 |
| SAT attenuation | -99 (-103, -95) | -98 (-101, -94) | -99 (-102, -95) | <0.001 |
| VAT volume index | 466 (319, 640) | 494 (349, 660) | 402 (278, 554) | <0.001 |
| VAT attenuation | -86 (-90, -80) | -86 (-90, -82) | -87 (-91, -82) | <0.001 |
| EAT volume index | 52 (36, 72) | 52 (37, 70) | 43 (32, 59) | <0.001 |
| EAT attenuation | -63 (-69, -55) | -61 (-69, -55) | -57 (-65, -53) | <0.001 |
| IMAT volume index | 107 (71, 163) | 106 (70, 160) | 89 (59, 137) | <0.001 |
| IMAT attenuation | -71 (-76, -67) | -71 (-75, -67) | -71 (-75, -67) | 0.004 |
| Total adipose tissue | 1632 (1123, 2212) | 1692 (1190, 2194) | 1312 (976, 1786) | <0.001 |
| Muscle to adipose ratio | 0.55 (0.41, 0.75) | 0.56 (0.43, 0.74) | 0.72 (0.55, 0.97) | <0.001 |
| Bone volume index | 349 (311, 387) | 364 (328, 401) | 341 (304, 378) | <0.001 |
| Bone attenuation | 255 (226, 285) | 252 (224, 281) | 265 (241, 291) | <0.001 |

Supplemental Table 1: Body composition measures as a function of stress modality and exercise stress duration in the combined internal and external SPECT populations. Values are presented as median (interquartile range). EAT – epicardial adipose tissue, IMAT – intermuscular adipose tissue, SAT – subcutaneous adipose tissue, SM – skeletal muscle, VAT – visceral adipose tissue.

Supplemental Table 2:

|  | Internal SPECT | External SPECT |
| --- | --- | --- |
|  | AUC (95% CI) | AUC (95% CI)) |
| Combined Body Composition | 0.756 (0.731 - 0.781) | 0.771 (0.752 - 0.789) |
| Age | 0.658 (0.629 - 0.686) | 0.717 (0.698 - 0.737) |
| BMI | 0.611 (0.580 - 0.641) | 0.583 (0.561 - 0.605) |
| SM volume index | 0.574 (0.544 - 0.604) | 0.647 (0.625 - 0.668) |
| SM attenuation | 0.727 (0.700 - 0.753) | 0.760 (0.742 - 0.779) |
| SAT volume index | 0.319 (0.290 - 0.348) | 0.339 (0.318 - 0.360) |
| SAT attenuation | 0.573 (0.543 - 0.602) | 0.574 (0.551 - 0.596) |
| VAT volume index | 0.431 (0.401 - 0.461) | 0.466 (0.443 - 0.488) |
| VAT attenuation | 0.505 (0.476 - 0.534) | 0.452 (0.430 - 0.475) |
| EAT volume index | 0.431 (0.400 - 0.460) | 0.385 (0.364 - 0.407) |
| EAT attenuation | 0.580 (0.550 - 0.610) | 0.593 (0.571 - 0.615) |
| IMAT volume index | 0.453 (0.424 - 0.483) | 0.458 (0.435 - 0.480) |
| IMAT attenuation | 0.564 (0.535 - 0.594) | 0.486 (0.463 - 0.508) |
| Total adipose tissue | 0.674 (0.645 - 0.704) | 0.656 (0.635 - 0.677) |
| Muscle to adipose ratio | 0.702 (0.674 - 0.729) | 0.706 (0.686 - 0.726) |
| Bone volume index | 0.514 (0.483 - 0.544) | 0.552 (0.529 - 0.574) |
| Bone attenuation | 0.562 (0.531 - 0.593) | 0.610 (0.588 - 0.632) |
| Combined Body Composition + Age | 0.795 (0.771 – 0.819) | 0.804 (0.787 – 0.821) |

Supplemental Table 2: Area under the receiver operating characteristic curve (AUC) for predicting exercise stress duration of at least 7 minutes in patients undergoing exercise stress. Age and BMI were modeled as inverse measures to simplify comparisons to the composite body composition score. CI – confidence interval, EAT – epicardial adipose tissue, IMAT – intermuscular adipose tissue, SAT – subcutaneous adipose tissue, SM – skeletal muscle, SPECT – single photon emission computed tomography, VAT – visceral adipose tissue

|  | Overall PET  (N=26553) | “Unfit” Body composition  n=21387 | “Fit” Body composition  n=5166 | p-value |
| --- | --- | --- | --- | --- |
| Age, median (IQR) | 67 (58, 75) | 69 (60, 76) | 60 (51, 69) | <0.001 |
| Male, n (%) | 15711 (59.2%) | 12992 (60.7%) | 2719 (52.6%) | <0.001 |
| BMI, median (IQR) | 29.5 (25.6, 34.8) | 30.4 (26.3, 36.3) | 26.4 (23.6, 29.8) | <0.001 |
| Past medical history, n (%) |  |  |  |  |
| Hypertension | 20836 (78.9%) | 17432 (81.7%) | 3404 (67.1%) | <0.001 |
| Diabetes mellitus | 9779 (36.9%) | 8550 (40.0%) | 1229 (23.8%) | <0.001 |
| Dyslipidemia | 19283 (73.0%) | 16021 (75.1%) | 3262 (64.3%) | <0.001 |
| Family history | 6807 (26.0%) | 5571 (26.3%) | 1236 (24.4%) | 0.006 |
| Smoking | 4840 (18.3%) | 4039 (18.9%) | 801 (15.8%) | <0.001 |
| Prior CAD | 8241 (31.0%) | 6764 (31.6%) | 1477 (28.6%) | <0.001 |
| Imaging results |  |  |  |  |
| Stress TPD | 4.7 (1.7, 11.1) | 4.8 (1.8, 11.9) | 4.3 (1.5, 10.8) | <0.001 |
| Rest TPD | 0.8 (0.1, 3.7) | 0.8 (0.1, 3.7) | 0.8 (0.0, 3.9) | 0.13 |
| Stress LVEF | 67.5 (56.0, 75.4) | 67.3 (55.6, 75.4) | 67.9 (57.6, 75.2) | 0.005 |
| Coronary artery calcium | 143 (0, 1002) | 198 (0, 1155) | 29 (0, 433) | <0.001 |
| Myocardial flow reserve | 2.31 (1.81, 2.90) | 2.26 (1.77, 2.82) | 2.60 (2.01, 3.25) | <0.001 |

Supplemental Table 3: Population characteristics of the PET population. BMI - body mass index, CAD - coronary artery disease, IQR - interquartile range, LVEF – left ventricular ejection fraction, TPD – total perfusion deficit.

Supplemental Figure 1:

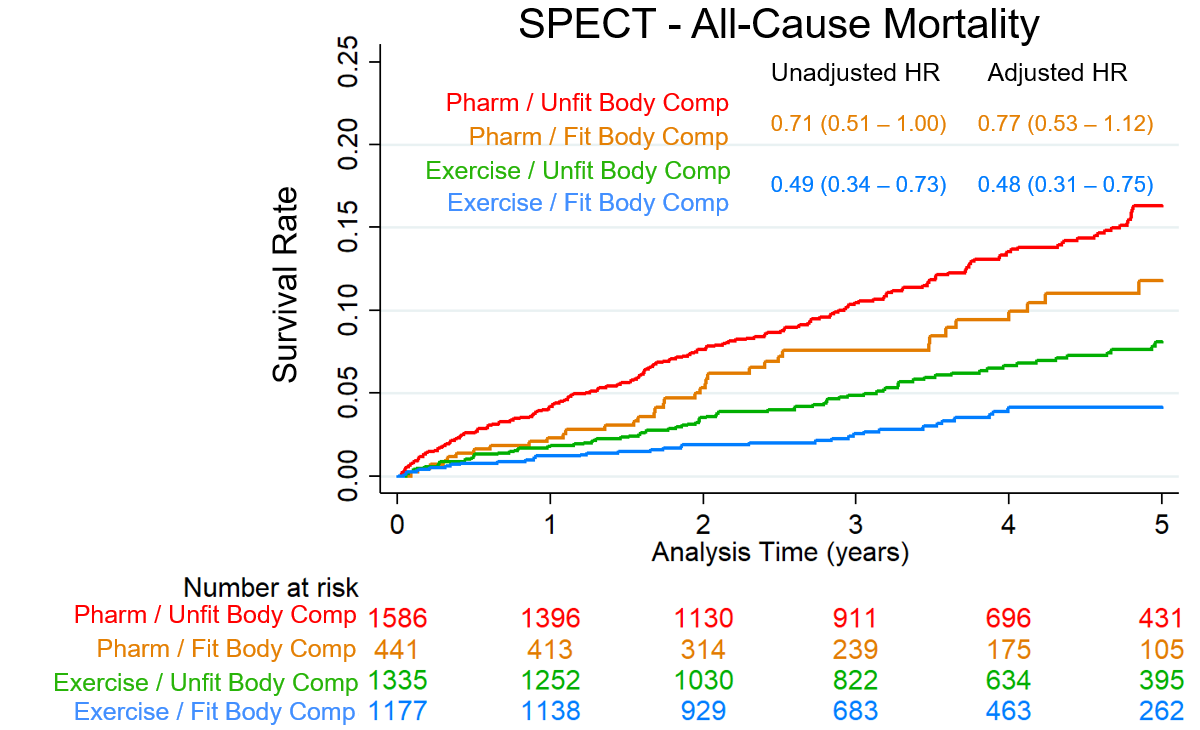

Supplemental Figure 1: Kaplan-Meier survival curves stratified by mode of stress and body composition in the external population. Mode of stress included exercise or pharmacologic (pharm). The body composition “fitness” score was derived from a logistic regression model in patients undergoing exercise stress in the internal population (n=2512), with thresholds also derived in the internal population. Hazard ratios (HR) reflect the risk associated with high body composition “fitness” score among patients undergoing pharmacologic or exercise stress. Multivariable models include age, sex, medical history, stress total perfusion deficit, left ventricular ejection fraction, and coronary artery calcium.
